# Using telehealth consultations for healthcare provision to patients from racial/ethnic minorities: A systematic review

**DOI:** 10.1101/2021.08.25.21262592

**Authors:** Mandy Truong, Ladan Yeganeh, Olivia Cook, Kimberley Crawford, Pauline Wong, Jacqueline Allen

**Affiliations:** Monash Nursing and Midwifery, Monash University, Clayton, Victoria, Australia; Menzies School of Health Research, Northern Territory, Australia

**Author notes:** Corresponding author: Mandy Truong, Monash Nursing and Midwifery, Monash University, Level 3, Building 13D, 35 Rainforest Walk, Clayton 3800, Australia;.

**Keywords:** telehealth, telemedicine, minority health, systematic review, racial/ethnic minorities

## Abstract

**Objective:** The COVID-19 pandemic has seen a rapid adoption of telehealth consultations, potentially creating new barriers to healthcare access for racial/ethnic minorities. This systematic review explored the use of telehealth consultations for people from racial/ethnic minority populations in relation to health outcomes, access to care, implementation facilitators and barriers, and satisfaction with care.

**Materials and Methods:** This review followed the Preferred Reporting Items for Systematic Reviews and Meta-Analysis and the JBI Manual for Evidence Synthesis. Five major databases were searched to identify relevant studies. Screening, full-text review, quality appraisal and data extraction were all completed independently and in duplicate. A convergent integrated approach to data synthesis was applied with findings reported narratively.

**Results:** A total of 28 studies met the inclusion criteria. Telehealth-delivered interventions were mostly effective for the treatment/management of physical and mental health conditions including depression, diabetes and hypertension. In several studies, telehealth improved access to care by providing financial and time benefits to patients. Technological difficulties were the main barriers to effective telehealth consultation, although overall satisfaction with telehealth-delivered care was high.

**Discussion:** Telehealth-delivered care for racial/ethnic minorities offers promise across a range of conditions and outcomes, particularly when delivered in the patient’s preferred language. However, telehealth may be problematic for some due to cost and limited digital and health literacy.

**Conclusion:** The development and implementation of guidelines, policies and practices in relation to telehealth consultations for racial/ethnic minorities should consider the barriers and facilitators identified in this review to ensure existing health disparities are not exacerbated.

## BACKGROUND AND SIGNIFICANCE

The global COVID-19 pandemic has seen a rapid adoption of telehealth in lieu of face-to-face consultations due to the need for social distancing and minimization of patient and healthcare provider physical contact.[1–3] As a result, the use of telehealth to deliver healthcare services has become widespread.[4] Implementation of this fast transition has been ad hoc with little evidence to guide such widespread adoption. Consequently, there is a risk that quality of care could be compromised and new access barriers may emerge for underserved groups such as racial and ethnic minorities who already experience health disparities.[5 6]

Telehealth consultations, including telemedicine, involve the use of telecommunication technologies between healthcare providers from any healthcare or social care discipline and patients in real time to transmit voice, images and data for healthcare and health education.[7] Using telehealth consultations, healthcare providers can deliver healthcare services and information directly to patients located elsewhere, addressing access problems related to distance and transport, waiting times, cost and limited patient and clinician time.[7–10] Pre-COVID-19, telehealth consultations in Western countries were largely limited to rural and remote patients, psychiatry or niche subspecialties such as cancer genetics.[11–14]

Existing systematic reviews of telehealth consultations have been conducted in a variety of clinical settings with generally positive findings in relation to patient satisfaction outcomes and consultation effectiveness.[15–17] In a review of quantitative studies, healthcare delivered by videoconferencing was effective in assessing patients’ health conditions and improving patients’ health conditions in 98% of the included studies.[18] This review also found that telehealth consultations improved treatment compliance and accountability for some participants.[18] Additionally, telehealth services have been found to improve access to care, and communication and engagement with clinicians from patients’ and informal carers’ perspectives.[16] However, telehealth services were not an effective means of psychosocial support in some studies and poor audio-visual quality limited patients’ satisfaction with healthcare.[16]

Patients and communities from racial/ethnic minorities are at risk of poorer health outcomes than the majority population because of structural inequities, including lack of access to health information in languages other than English, underutilization of interpreter services, lack of culturally appropriate services, and racism and discrimination.[19–23] The use of telehealth consultations may further complicate healthcare access for patients from racial/ethnic minority backgrounds due to their limited health and digital literacy, and their challenges in navigating mainstream healthcare systems.[23 24] These access barriers are potentially heightened during the COVID-19 pandemic.[7]

Despite previous reviews of telehealth effectiveness for different patient populations such as older populations and rural and remote communities, no reviews have been undertaken of studies assessing telehealth consultations specifically for patients and communities from racial/ethnic minority backgrounds. An evidence synthesis focused on patients and communities from racial/ethnic minority groups would generate an evidence base for the tailoring of telehealth consultations to their diverse needs.

## OBJECTIVE

The objective of this review was to explore the health outcomes, implementation facilitators and barriers, and satisfaction with care, in relation to telehealth consultations for racial/ethnic minority populations in order to make recommendations for practice and future research.

## MATERIALS AND METHODS

### Information sources and search strategy

The protocol for this systematic review was registered in the international register of systematic reviews PROSPERO (CRD 42020221017) and was conducted in accordance with the PRISMA guidelines.[25] Studies were identified from a search of the following 5 electronic databases: Ovid Medline, Ovid PsycINFO, EMBASE, and CINAHL via EbscoHOST and Scopus platforms. The search was limited to studies published in English between 1 January 2005 and 9 October 2020. Additionally, a search for existing systematic reviews on the same topic was conducted in: all EBM reviews via Ovid, Cochrane Database of Systematic Reviews, Joanna Briggs Institute, and PROSPERO. Reference lists of included studies were hand-searched for relevant studies.

A list of terms was formulated for the concepts: “telehealth” and “racial or ethnic minority”. These terms were mapped to subject headings (e.g., Medical Subject Headings or equivalent) and keywords. Boolean operators were used to group subject headings and keywords to create a search strategy (Supplementary file X).

### Eligibility Criteria

To be included in the review the study had to meet the following inclusion criteria:

a. Sample population included patients from racial/ethnic minorities, their carers or healthcare staff who provided care to racial/ethnic minorities, with study findings disaggregated by racial/ethnic minority groups.
b. For this review, we describe the intervention as ‘telehealth consultation’. Telehealth consultations occurred instead of a face-to-face consultation to provide a health service such as a clinical assessment or diagnosis and/or management of a health condition (physical or mental). Support services provided as standard practice (e.g., phone interpreting, telemonitoring without direct interaction with a healthcare provider) were excluded. Studies that primarily focussed on delivery of health promotion programs or health-related education via telehealth were also excluded.
c. Included telehealth consultations in healthcare settings (e.g., hospitals, primary care clinics)
d. Reported individual health outcomes (e.g., physical health, mental health). Individual level outcomes could be related to patients/health consumers and care-givers.
e. Reported health service outcomes (e.g., readmission rates, barriers to implementation). Health service level outcomes may be related to access to care, service delivery, satisfaction with care and cost-effectiveness
f. Used quantitative, qualitative or mixed methods study designs
g. Published in a peer-reviewed journal

Reviews of telehealth consultations and healthcare for Indigenous peoples have been published.[26–30] We therefore did not include Indigenous peoples in the current review. Publications were excluded if they were opinion pieces, systematic reviews and meta-analyses, conference abstracts or proceedings or duplicate publications using the same sample and reporting the same outcomes.

### Data screening and extraction

Search results were exported from the electronic databases into the systematic review management platform Covidence (Veritas Health Innovation, 2021). Screening was undertaken in two phases. Following the removal of duplicates, studies were screened independently in duplicate by two reviewers on title and abstract. Conflicts were resolved by a third reviewer. In the second screening phase, full text articles were screened independently in duplicate by two reviewers and conflicts were resolved by a third reviewer.

Data extraction was performed by two reviewers using Covidence, with differences resolved by a third reviewer. Extracted data comprised the characteristics of each study: country of study, health setting, study aim, study design, sample size and participants’ characteristics (e.g., gender, age, race/ethnicity/cultural background), health conditions, and the details of the telehealth intervention. Data were also extracted pertaining to study findings: health outcomes (e.g., blood pressure, depression symptoms) and health access and delivery outcomes (e.g., satisfaction with care, implementation barriers and facilitators).

### Methodological quality assessment of studies

Risk of bias and critical appraisal were conducted independently in duplicate by two reviewers using the Joanna Briggs Institute (JBI) Critical Appraisal tools.[31] The JBI Critical Appraisal Tools assess the methodological quality of studies of various design and the extent to which the risk of bias is addressed by authors. For each criterion assessed as ‘met’ on the appropriate tool, a score of ‘1’ was applied. Where criteria were assessed as ‘not met’ or if it was ‘unclear’ to the reviewer, a score of ‘0’ was applied. The scores were then calculated for each study and converted to a final quality rating of ‘low’, ‘moderate’ or ‘high’. The following JBI Critical Appraisal Tools and scoring parameters were implemented in this review: Checklist for randomized-controlled trials (RCTs) (score out of 10; Low 0-3, Moderate 4-7, High 8-10); the Checklist for quasi-experimental studies (score out of 9; Low 0-3, Moderate 4-6, High 7-9); Checklist for Analytical Cross-Sectional Studies (score out of 8; Low 0-3, Moderate 4-6, High 7-8); the Checklist for Qualitative Research (score out of 10; Low 0-3, Moderate 4-7, High 8-10); the Checklist for Cohort Studies (score out of 11; Low 0-4, Moderate 5-8, High 9-11); and, the Checklist for Case Series Studies (score out of 10; Low 0-3, Moderate 4-7, High 8-10). (See Supplementary file for critical appraisal criteria for each study design.)

For mixed methods studies, we used two questions screening questions and five questions from the ‘mixed methods studies’ section of the Mixed Methods Appraisal Tool version 2018 (MMAT),[32] (score out of 7; Low 0-2, Moderate 3-5, High 6-7) which are related to the rationale for using a mixed methods study design and level of interpretation and integration of the qualitative and quantitative components of the study.[32]

### Synthesis of studies

Extracted data were exported from Covidence into a Microsoft Excel spreadsheet for analysis. A convergent integrated approach to data synthesis was taken in accordance with the JBI methodology for mixed methods systematic reviews.[31] Due to the heterogeneity of study methodology, design, sample characteristics, and outcome measures, there was no opportunity to conduct a meta-analysis of quantitative studies nor a meta-synthesis of qualitative studies. A narrative synthesis was thus conducted where both qualitative and quantitative data were tabulated together and categorized according to the four outcomes of interest: health outcomes, health access, implementation barriers and facilitators to telehealth consultations, and satisfaction with care. A comparison between studies reporting on the same outcome was performed, with similar and divergent findings reported. Study characteristics such as country and setting, study methodology and design, sample sizes and participant characteristics, were examined by calculating frequencies and proportions.

## RESULTS

### Study selection

The combined searches from the four databases identified 3,314 unique articles which were screened using the inclusion and exclusion criteria. A total of 28 articles met the inclusion criteria for the systematic review (Figure 1).

**Figure 1:**
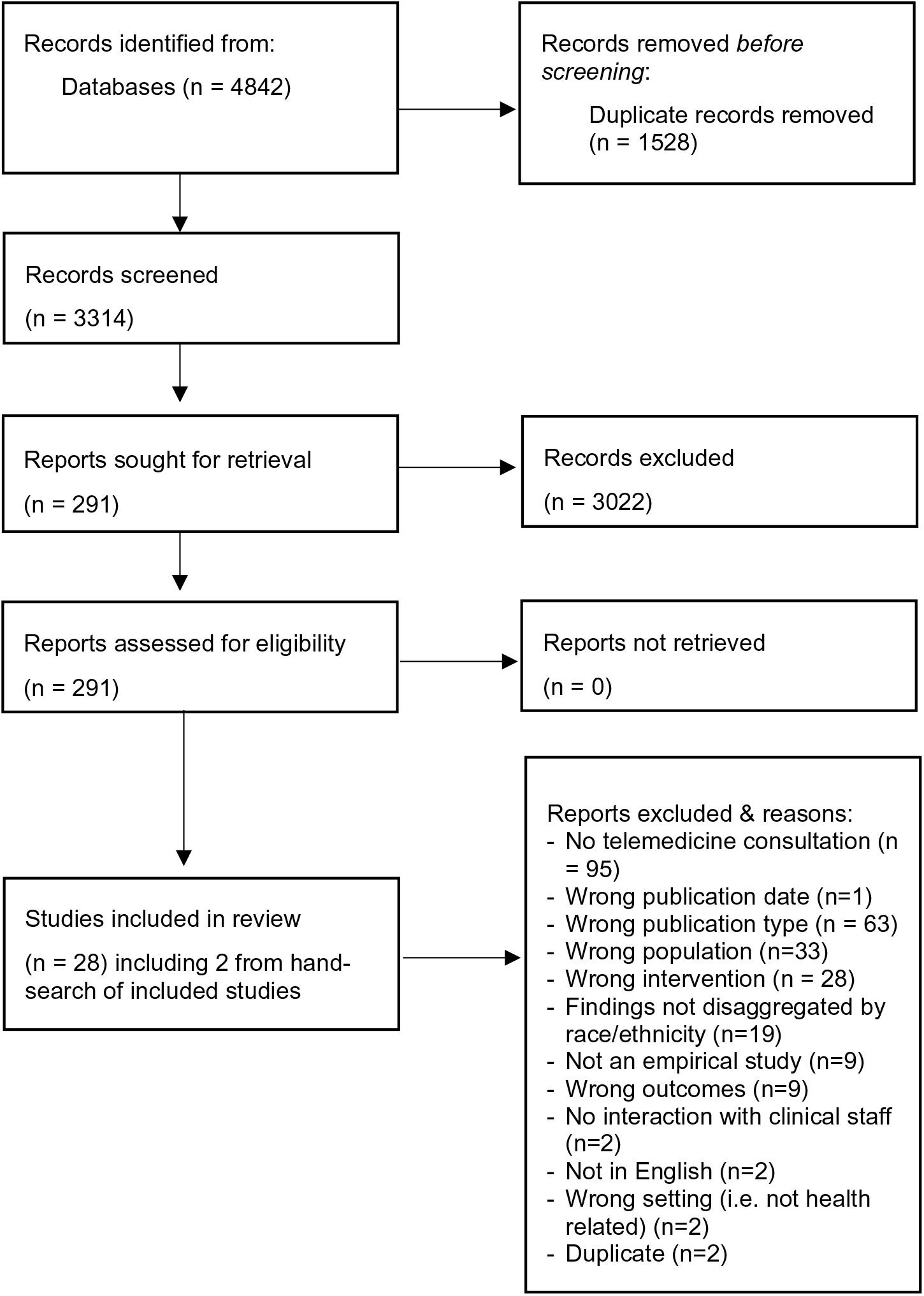
Flow diagram of study selection. Adapted from PRISMA flow-chart [25]

### Study characteristics

Studies were published between 2005 and 2019 and comprised 4,301 patients and 12 caregivers from racial/ethnic minorities, and 34 healthcare practitioners. The majority of studies (n=23, 82.1%) examined groups located in the United States and utilized quantitative study designs (n=19, 67.9%). The most common health conditions examined were mental health conditions (n=17, 60.7%), of which nine focused on depression. The most frequent study setting was community health centers (n=8, 28.6%), followed by primary health clinics (n=5, 17.9%), psychiatric or trauma treatment centers (n=4, 14.3%), the community (n=3, 10.7%) and HIV clinics (n=2, 7.1%). The racial/ethnic minority populations included in the studies were predominantly Latino/a or Hispanic (n=13, 46.4%) and African American (n=7, 25.0%). Seven studies included Asian participants from Korean and Chinese communities, three studies included participants from refugee and immigrant backgrounds and one study described their minority population as ‘non-white ethnic group’. Characteristics of included studies are presented in Table 1. (Supplementary file Table 1 presents detailed information related to study characteristics.)

**Table 1:** Summary of included telehealth articles (n=28)

|  |  | n | % |
| --- | --- | --- | --- |
| **Study location** | United States | 23 | 82.1 |
|  | Australia | 1 | 3.6 |
|  | Denmark | 1 | 3.6 |
|  | Denmark & Sweden | 1 | 3.6 |
|  | Korea, Vietnam, Cambodia & Uzbekistan | 1 | 3.6 |
|  | United Kingdom | 1 | 3.6 |
| **Year published** | 2005-2009 | 3 | 10.7 |
|  | 2010-2014 | 16 | 57.1 |
|  | 2015-2019 | 9 | 32.1 |
| **Study design** | Randomized controlled trial | 11 | 39.3 |
|  | Cohort | 1 | 3.6 |
|  | Quasi-experimental | 2 | 7.1 |
|  | Cross-sectional | 4 | 14.3 |
|  | Case series | 1 | 3.6 |
|  | Mixed methods | 6 | 21.4 |
|  | Qualitative | 3 | 10.7 |
| **Study setting** | Community health center | 8 | 28.6 |
|  | Primary care clinic | 5 | 17.9 |
|  | Psychiatric or trauma treatment center | 4 | 14.3 |
|  | Community | 3 | 10.7 |
|  | HIV clinic | 2 | 7.1 |
|  | Other | 6 | 21.4 |
| **Health condition <sup>a</sup>** | Mental health | 17 | 60.7 |
|  | Cardiovascular related (e.g., hypertension) | 8 | 28.6 |
|  | Diabetes | 5 | 17.9 |
|  | Infectious diseases (e.g., HIV, Hepatitis C) | 2 | 7.1 |
|  | Primary care | 1 | 3.6 |
|  | Health counselling | 1 | 3.6 |
| **Racial/ethnic background of minority participants <sup>b</sup>** | Hispanic/Latino/a | 13 | 46.4 |
|  | African American | 7 | 25.0 |
|  | Korean & Korean American | 5 | 17.9 |
|  | Chinese American | 2 | 7.1 |
|  | Other (e.g., refugees, non-white ethnic group) | 4 | 14.3 |

Percentages may not add up to 100 due to rounding
<sup>a</sup> Some studies included more than one health condition
<sup>b</sup> Some studies included participants from more than one racial/ethnic group

### Quality assessment

The quality of the included studies was mixed (see Supplementary file). All 11 RCTs reported random sampling from patients presenting for clinical care. There was variation in the reporting of allocation concealment and differences in baseline characteristics between groups in some RCTs, however most reported completion of follow-up assessments and an intent to treat approach to data analysis. As all RCT studies investigated a telehealth consultation between a practitioner and a patient, blinding of participant and practitioner was not possible. Accordingly, no RCT studies reported blinding of participants to intervention group or blinding of practitioners delivering the intervention to group allocation. The number of quality indicators for RCT studies reported ranged from 3/10 to 7/10. Each of the two quasi-experimental studies employed a pre/post design and there were mixed results regarding study quality. The quality of reporting for the one study utilizing a cohort design and for the one study using a case series design was high. Overall, the quality of reporting for studies using cross-sectional, mixed methods and qualitative designs was low to medium.

### Telehealth consultations

Data pertaining to telehealth consultations are presented in Table 2. Across the included studies, telehealth consultations were primarily mental health consultations (n=17, 61%) such as cognitive behavioral therapy (CBT) or psychiatry (telepsychiatry). Physical health consultations were related to chronic disease management (n=9, 32.1%) and clinical monitoring (n=2, 7.1%). Physical health diseases that were managed by telehealth consultations included diabetes (n=2, 7%), hypertension (n=3, 11%) and infectious diseases (n=2, 7%). Telehealth consultations were used to monitor patients’ clinical status in two studies. One study used a telephone triage system and computer-supported decision-making software.[33] The other study monitored patient’s vital signs and other health related information ‘remotely’; and this was then used to inform virtual ‘wellness’ visits by a nurse practitioner who discussed the patient’s medications and recent health data.[34]

**Table 2.** Characteristics of included studies by study design.

| First author, year | Country | Health condition | Sample size and participant race/ethnicity | Telemedicine intervention |
| --- | --- | --- | --- | --- |
| **Randomized controlled trial** |  |  |  |  |
| Alcantara 2016 [60] | USA | Mental health (Depression) | Total n=257; Latino<br>Usual care n=86<br>Face-to-face ECLA=84<br>Telephone ECLA=87 | Telephone plus care management<br>CBT<br>6–8 weekly/bi-weekly sessions |
| Chong 2012 [43] | USA | Mental health (Depression) | Overall n=167; Hispanic<br>Usual care n=87<br>Intervention=80 | Telepsychiatry consultations |
| Dwight-Johnson 2011 [53] | USA | Mental health (Depression) | Total n=101; Latino<br>Usual care n=52<br>Intervention n=50 | Telephone CBT<br>8 sessions |
| Han 2010 [36] | USA | Hypertension | Total n=360; Korean<br>More intensive intervention<br>telephone counseling<br>(MIC)=182<br>Less intensive intervention<br>telephone counselling<br>(LIC)=178 | Telephone counseling<br>MIC= biweekly counseling<br>LIC= monthly counseling<br>12-month duration |
| Himelhoch 2013 [55] | USA | Mental health (Depression) | Total n=34; AA=32, AI=1,<br>other=1, Hispanic=2<br>Face-to-face CBT n=18<br>Telephone CBT n=16 | Telephone CBT<br>11-sessions |
| Jackson 2012 [37] | USA | Hypertension | Overall n=573; AA=284,<br>White=289<br>Number of participants in each<br>study arm not reported | Arm 1: HBPM with remote<br>medication management.<br>Arm 2: HBPM with behavioral<br>management.<br>Arm 3: Combined: HBPM,<br>medication management,<br>behavioral intervention<br>Arm 4: usual care |
| Kim 2011 [38] | USA | Hypertension | Total n=359; Korean<br>MIC n=182<br>LIC n=177 | Telephone counseling<br>MIC= biweekly, LIC= monthly<br>counseling<br>12 -month duration |
| Moreno 2012 [44] | USA | Mental health (Depression) | Overall n=182; Hispanic<br>Usual care n=87<br>Intervention=80 | Telepsychiatry consultations<br>Six monthly sessions over 3 years |
| Vahia 2015 [47] | USA | Mental health | Total n=27, Latino<br>In-person testing n=11<br>Telephone testing n=11 | Telepsychiatry for neuro-cognitive assessment |
| Warren 2015 [33] | United Kingdom | Primary care | Total n=12,132; White=11276,<br>other ethnic=476<br>Usual care =4093<br>GP triage n=4034<br>Nurse triage n=3704 | Telephone triage using computer-supported decision-making software by doctor or nurse. |
| Yeung 2016 [49] | USA | Mental health | Total n=190; Chinese<br>Usual care n=97<br>T-CSCT n=93 | Telepsychiatry consultations |
| **Cohort** |  |  |  |  |
| Berg 2009 [35] | USA | Diabetes | Total n=980; Hispanic<br>Usual care n=490<br>Intervention n=490 | Disease management diabetes program and scheduled nurse education sessions.<br>24-hour access to a nurse counselor. |
| **Quasi-experimental** |  |  |  |  |
| Jang 2014 [56] | USA | Mental health (Depression) | n=14; Korean | Video-telecounseling consultations<br>4 weekly sessions (30min in each) |
| Mayes 2010 [39] | USA | Diabetes | n=16, Hispanic | Video-telehealth consultations assisted by Promotoras and primary care physician connecting to a medical specialist |
| **Cross-sectional** |  |  |  |  |
| Mucic 2010 [45] | Denmark & Sweden | Mental health | n=61; refugees=45, asylum seekers=12, migrants=3, domestic=1 | Video-telepsychiatry consultations |
| Park 2019 [40] | Korea, Vietnam, Cambodia, Uzbekistan | Health counselling | n=442; Korean | Video-telehealth counseling |
| Schulz 2014 [42] | Australia | Infectious diseases | n=119; refugees<br>n=29 doctors | Video-telehealth consultations |
| Yeung 2009 [50] | USA | Mental health | n=9; Chinese | Video-telehealth consultations |
| **Case series** |  |  |  |  |
| Stewart 2017<sup>a</sup> [58] | USA | Mental health | n=4; Hispanic | Weekly TF-CBT sessions |
| **Mixed methods** |  |  |  |  |
| Bagchi 2018 [34] | USA | Diabetes, hypertension | n=10; AA | Video-telehealth consultations |
| Glueckauf 2012 [54] | USA | Mental health (Depression) | Total n=12; AA caregivers<br>Face-to-face CBT n=6<br>Telephone CBT n=6 | Telephone CBT<br>Twelve, 1-hr, weekly sessions |
| Mucic 2007 [51] | Denmark | Mental health | n=23; refugees | Video-telepsychiatry consultations |
| Stewart 2017<sup>b</sup> [59] | USA | Mental health | n=15; Hispanic=7, AA=6, White=2 | TF-CBT via video<br>Weekly sessions (range 12-19 in total) |
| Uebelacker 2011 [46] | USA | Mental health (Depression) | Total n=38; Latino<br>Usual care n=19<br>Intervention n=19 | Telehealth depression care management |
| Ye 2012 [48] | USA | Mental health | n=16; Korean | Video-telehealth consultations |
| **Qualitative** |  |  |  |  |
| Aisenberg 2012 [52] | USA | Mental health (Depression) | Overall n=106; Latino<br>Usual care n=51<br>Intervention n= 50<br>Physicians n=5 | Telephone CBT<br>Eight-sessions |
| Pekmezaris 2016 [41] | USA | Heart failure | n=4; AA, Hispanic | Video-telemonitoring consultations<br>Weekly for 3 months |
| Saberi 2013 [57] | USA | HIV | n=14; AA | Video-telehealth consultations |

AA=African American, AI=American Indian
CBT= Cognitive Behavioral Therapy
ECLA= Engagement and Counseling for Latinos intervention
HIV= Human Immunodeficiency Virus
HBPM=Home Blood Pressure Monitoring
T-CSCT=Telepsychiatry-based Culturally Sensitive Collaborative Treatment
TF-CBT=Trauma Focused - Cognitive Behavioral Therapy

The telehealth consultations and interventions were delivered by a range of healthcare professionals including: nurses (n=10, 35.7%),[33–42] physicians (n=13, 46.4%)[33 37 39 40 42–51] including psychiatrists, primary care physicians and specialists, allied health professionals (n=9, 32.0%)[46 52–59] such as social workers, counsellors and psychologists, and paraprofessional outreach workers in one study.[39] In eighteen studies (64%), the healthcare professionals delivering the telehealth intervention were bilingual. The mode used to deliver the telemedicine intervention was video conferencing in 16 studies (57.1%) and telephone in 11 studies (39.3%); one study used both internet video and telephone communication.[39]

### Health outcomes

In total, 14 studies measured health outcomes with evidence of effectiveness of telemedicine on measures of mental health, cardiovascular health and diabetes related outcomes. Three RCTs focused on health outcomes related to treatment of depression. These three studies assessed the effect of a telehealth CBT intervention on depression severity compared with face-to-face CBT[55 60] or compared with anti-depressant therapy.[53] Among Latino patients, worry symptoms were reduced[60] and depression symptoms were improved[55] following the use of culturally adapted telephone-based CBT compared to face-to-face CBT. In the study by Dwight-Johnson[53], Latino patients receiving culturally adapted telephone-based CBT showed greater improvement in depression symptoms over 6 months compared to patients who received usual care with anti-depressant medications.

The effects of telepsychiatry consultations or remote management on improving depression outcomes were reported in three RCTs.[43 46 49>] In a study of Hispanic patients with depression, monthly telepsychiatry consultations and usual care were equally effective in reducing depression scores after 6 months.[43] In another pilot RCT [46], telephone-based depression care management plus usual care was compared with the usual care only group. The findings of this study showed a trend for lower levels of depression in the intervention group over time compared with the control group.[46] In another study, the effectiveness of culturally sensitive telepsychiatry treatment and care management was compared to usual care for Chinese Americans with depression.[49] This study found greater improvement in depressive symptoms following telepsychiatry compared with usual care.[49] In a single group pre-test post-test design with Korean immigrants, weekly sessions of telepsychiatry for four weeks significantly reduced depression scores at immediate follow-up and at three-months follow-up.[56] In a pilot study using a pre-post two group design focusing on African American caregivers of older adults with dementia, telephone-based and face-to-face CBT interventions were equally effective in reducing depression scores.[54]

Three studies assessed the effects of telemedicine on cardiovascular and diabetes health outcomes.[37–39] In one RCT study of patients with hypertension from Korea, bi-weekly and monthly hypertension-related telephone counselling, and 12-month home monitoring of BP were effective in improving long-term BP outcomes[38]. The second study employing a RCT design assessed the effectiveness of a telemedicine intervention emphasizing medication management and home BP monitoring with African American patients living with hypertension [37]. Jackson et al.[37] found significant improvement in mean systolic blood pressure at 12-months and at 18-months following the intervention compared with usual care. A study by Mayes [39] assessed the impact of telemedicine on blood glucose regulation among Hispanic patients living with type 2 diabetes and found that telemedicine monitoring significantly improved participants’ blood glucose regulation.

### Barriers and facilitators

Three studies identified technology as a barrier to the implementation of telehealth, which was not unique to a health condition or study design. A mixed method study involving African Americans and Hispanics with mental health conditions identified initial issues with logging into the videoconferencing software and technical problems with the equipment.[59] A cross sectional study involving refugees with infectious diseases experienced technical difficulties in a quarter of the first telehealth consultations, although this significantly improved with experience.[42] A qualitative study involving African Americans and Hispanics with heart failure found that there were initial concerns with using the equipment, but these dissipated with use.[41] This study also reported issues with internet connection, using the equipment, patients managing the intervention alone at home, and concerns with patient literacy. The combined use of audio and text in the telemonitoring program assisted patients’ understanding.[41]

Six studies identified facilitators to telehealth implementation. The facilitators were not unique to a health condition or race/ethnic background. Telehealth consultations using video conferencing supported health professionals to provide education to patients with mental health conditions by sharing PowerPoint presentations and other written material.[58 59] Telehealth permitted caregiver engagement and could be delivered in a language of the patient and caregiver’s preference.[58 59] Patients from ethnic minorities reported ease of access to medical help through the GP telephone triage system.[33] Telehealth allowed patients with HIV quick access to their providers and increased their likelihood of attending appointments.[57] Patients with diabetes and hypertension gave favorable ratings to the accessibility of telehealth.[34] Strategies to assist with telehealth consultations included: face-to-face meetings prior to establishing telehealth to build relationships[59] and use of a

### Satisfaction with telehealth consultations

Overall, 16 studies (57.1%) explored satisfaction with telehealth consultations. Of these, nine were quantitative studies[33 40 43 45 50 53 55 56 60], five studies were mixed methods [34 46 48 51 59] and two studies used qualitative methods.[52 57] Results across the sixteen studies were mixed, with the majority of studies (n=11, 68.8%) reporting high levels of satisfaction with telehealth among patients, carers and health professionals.[34 40 43 45 50–53 56 57 59] Reasons for satisfaction with telehealth consultations included increased efficiency and convenience, enhanced privacy and reduced need for travel compared to in person face-to-face consultations. Some studies found no discernible differences in levels of satisfaction between telehealth groups and comparison groups (i.e. usual care or face-to-face interventions)[46 55 60] and participants in one study gave mixed feedback about telepsychiatry.[48] In a study examining telephone triage, patients from ethnic minorities reported higher satisfaction in the GP triage arm compared to usual care but lower satisfaction compared with white patients overall.[33]

## DISCUSSION

This systematic review examined the existing empirical literature on telehealth consultations related to racial/ethnic minorities. Findings from this review indicate that telehealth consultations for mental health and some physical health conditions between health practitioners and patients from racial/ethnic minorities offer promise across a range of outcomes and healthcare settings and can result in high levels of patient satisfaction. However, it is yet to be determined whether this translates to other healthcare settings, different health conditions and other racial/ethnic minority populations (particularly in countries outside the United States).

Despite the overall positive impact of telehealth consultations for patients from racial/ethnic minority backgrounds, some barriers and challenges were identified by the studies. Information technology required to implement telehealth consultations may be problematic for some patients from racial/ethnic minorities because of the cost of equipment, limited understanding of the use of equipment, and limited health literacy. This concurs with findings from previous reviews of telehealth consultations.[15 16 18] Additionally, there may be unique and unanticipated challenges to telehealth consultation delivery for some ethnic communities. Some racial and ethnic communities may experience anxiety regarding telecommunications including telehealth because of their perceived risk of fraudulent use of telecommunication or because of their fear of contact from immigration authorities.[46] Any barriers and challenges to use of telehealth consultations are likely to be further exacerbated by factors related to English language proficiency, cultural factors and lack of familiarity with mainstream health systems. This includes awareness of changes in government subsidies related to medical care and advice delivered via telehealth.[61]

Other literature notes that limited English proficiency can be a significant barrier to health service access and utilization for racial/ethnic minorities, including via telehealth consultations. A recent survey by Rodriguez et al.[62] found that patients with limited English proficiency had lower rates of telehealth consultation use compared with proficient English speakers. In English speaking countries, patients with limited English language proficiency may experience substantial difficulties with the use of telehealth and information technology equipment suggesting that these patients are at greater risk of receiving poorer quality healthcare when engaging in telehealth.

Furthermore, providing culturally appropriate healthcare is also important for racial/ethnic minorities.[63] In our review, some of the studies included culturally adapted materials in their telehealth consultations and used bilingual staff or interpreters and is likely to have contributed to the overall positive findings across the included studies. The cultural appropriateness and acceptability of telehealth consultations and development of cultural frameworks to guide telehealth use should be considered more broadly, particularly if services are being delivered transnationally.[64 65]

Overcoming barriers and challenges to telehealth should address the need for digital literacy and linguistically appropriate online information. Although the use of bilingual staff or interpreters to deliver a telehealth consultation may mitigate some of the challenges experienced by racial/ethnic minorities when using telehealth, patients may still face significant barriers due to poor digital literacy skills. Digital literacy refers to a person’s ability to use digital tools (e.g., health applications, patient portals, appointment booking) to access, understand and analyse information and communicate with others.[66 67] Previous studies suggest low digital literacy to be a contributing factor to racial/ethnic disparities among telehealth users.[68 69]

### Limitations of the current evidence-base and areas for further research

This review identified key limitations of existing research in telehealth for racial/ethnic minorities. Given the increasing and more widespread adoption of telehealth consultations across the health system due to the COVID-19 pandemic, it is important to understand its impact for patients from racial/ethnic minority groups to ensure equity of healthcare access and utilization. A recent study of telemedicine use during COVID-19 has already identified disparities in telemedicine access for African American patients compared to white patients.[6]

Further research is required in other areas such as acute and chronic disease management and in other care settings for racial/ethnic minorities such as cancer care, kidney disease, emergency department admissions, critical care, pediatrics and obstetrics/midwifery. Healthcare services and institutions should monitor use of telehealth consultations across patient demographics, including language and racial/ethnic background, to measure the impact on health equity. Although our search strategy identified studies conducted in non-English speaking countries, most studies were based in the United States of America. Additional studies are required in other countries with underserved populations who may have less access to information technologies.

Included studies were also limited by a lack of methodological rigor across most study designs, resulting in a significant gap in the current evidence base. Longitudinal studies are needed to determine whether, and to what extent, telehealth consultations affect health-related outcomes over time for racial/ethnic minorities. In particular, the effects of telehealth on early identification of disease and on the long-term management of chronic health conditions are required to maximize long-term health outcomes for racial/ethnic minorities.

Patients from marginalized racial/ethnic minority groups such as refugees were only specifically studied in three of the 28 studies.[42 45 51] Issues related to caregiver support and the optimal mode of telehealth consultation delivery (i.e., telephone vs videoconferencing) should also be explored in future research.

### Limitations of the review

Our review has several limitations. Only articles published in English were included therefore there is risk of publication bias. Additionally, the search period of 1 January 2005 to 9 October 2020, may have omitted earlier relevant studies. However, information technology enabling telehealth consultation has evolved considerably since the early 21^st^ century. We anticipate that studies published prior to 2005 would have limited relevance.

## CONCLUSION

This review has shown telehealth consultations to be mostly positive and beneficial to patients from racial/ethnic minority groups in terms of health outcomes, satisfaction with healthcare and accessibility of health services. Telehealth consultations delivered to patients in their preferred language or by bilingual health providers contributed to positive outcomes. Challenges to implementation of telehealth consultations across racial/ethnic minority populations were also identified and should be considered in the development and implementation of guidelines, policies and practices in relation to the use of telehealth consultations across healthcare. Further research is needed to understand the long-term impacts of telehealth use to ensure health disparities are not worsened.

## Data Availability

Data is available upon request to authors

## ACKNOWLEDGMENTS

None.

## SUPPLEMENTARY FILE

**Table 1.** Characteristics of included studies.

| First author, year | Country | Health condition and setting | Study aim | Sample size and participant race/ethnicity | Telemedicine intervention | Study findings | Quality appraisal score |
| --- | --- | --- | --- | --- | --- | --- | --- |
| **Randomized controlled trial** |  |  |  |  |  |  |  |
| Alcantara 2016 | USA | • Mental health (depression)<br>• Primary care clinics | • To examine whether Engagement and Counselling for Latinos (ECLA) reduced worry symptoms when compared to usual care and whether delivery modality (telephone vs. face-to-face) and patient sociodemographic characteristics moderated the effectiveness of ECLA on worry reduction. | • Total n=257<br>• All Latino<br>• Usual care n=86<br>• Face-to-face (ECLA) intervention n=84<br>• Telephone ECLA intervention n=87 | • Telephone plus care management CBT<br>• 6–8 weekly/bi-weekly sessions | • Participants in telephone ECLA experienced a statistically significant reduction in worry when compared to those in usual care and when compared to those in face-to-face ECLA | 5/10 |
| Chong 2012 | USA | • Mental health (depression)<br>• Community health center(s) | • To assess the feasibility and acceptability of telepsychiatry for low-income Hispanic patients with major depression | • Overall n=167<br>• All Hispanic<br>• Usual care n=87<br>• Intervention=80 | • Telepsychiatry consultations with bilingual providers | • Both intervention and usual care patients showed: a significant decrease in depression symptoms from intake to the 6-month follow-up and significant decrease in the number of days in which their symptoms rendered them less productive or worse.<br>• There was strong support for acceptability and less definite<br>support for the feasibility of telepsychiatry. | 7/10 |
| Dwight-Johnson 2011 | USA | • Mental health (depression)<br>• Community health center(s) | • To test the effectiveness of culturally tailored, telephone-based CBT for improving depression outcomes among Latino primary care patients | • Total n=101<br>• All Latino<br>• Usual care n=52<br>• Intervention n=50 | • Telephone CBT with bilingual providers<br>• 8 sessions | • Patients who received CBT by phone were more likely to experience improvement in depression scores over the six-month follow-up period compared with patients who received enhanced usual care. | 7/10 |
| Han 2010 | USA | • Hypertension<br>• Community | • To evaluate the use of bilingual nurse-delivered telephone counselling in Korean Americans to improve management of hypertension. | • Total n=360<br>• All Korean<br>• More intensive intervention telephone counseling n=182<br>• Less intensive intervention telephone counselling n=178 | • Telephone counseling with bilingual providers<br>• More intensive= biweekly counseling<br>• Less intensive group= monthly counseling<br>• 12-month duration | • Both groups showed improvement with regard to medication-taking, alcohol consumption and exercise but not smoking, with no significant group differences. | 5/10 |
| Himelhoch 2013 | USA | • Mental health (depression)<br>• HIV clinic(s) | • To pilot a randomized control trial of a telephone-based CBT intervention for low income, HIV infected | • Total n=34<br>• African American: 32 (94.1 %), American Indian | • Telephone CBT<br>• 11-sessions | • No statistically significant differences in depression treatment outcomes comparing face-to-face CBT to telephone CBT | 7/10 |
|  |  |  | African-Americans with major depression. | 1 (2.9 %), other 1 (2.9 %), Hispanic: 2 (5.7 %)<br>• Face-to-face CBT n=18<br>• Telephone CBT n=16 |  | • Telephone CBT improved adherence to HIV medication compared with face-to-face CBT. |  |
| Jackson 2012 | USA | • Hypertension<br>• Primary care clinics | • To examine whether there were differences in change in blood pressure (BP) for African American and non-Hispanic white patients in response to a medication management and tailored nurse-delivered telephone behavioral program | • Overall n=573<br>• African American n=284 (48.1%), non-Hispanic white n= 289 (49.1%)<br>• # of participants in each study arm not reported | • Study arm 1: Home BP monitoring with remote medication management.<br>• Study arm 2: Home BP monitoring with behavioral management including telephone health behavior modules.<br>• Study arm 3: Combined: home monitoring, triggered medication management, triggered behavioral intervention<br>• Study arm 4: usual care | • Improvement in mean systolic blood pressure post-baseline was greater for African American patients in the combined intervention, compared to African American patients in usual care, at 12 months and 18 months. | 7/10 |
| Kim 2011 | USA | • Hypertension<br>• Community | • To report the results of a clinical investigation to determine the sustainability of intervention effects to lower blood pressure (BP) through a short- term education via home telemonitoring of BP and regular counseling by bilingual nurses during 1 year | • Total n=359<br>• All Korean<br>• More intensive intervention telephone counseling (MIC) n=182<br>• Less intensive intervention telephone counselling (LIC) n=177 | • Telephone counseling with bilingual providers<br>• More intensive= biweekly counseling<br>• Less intensive group= monthly counseling<br>• 12-month duration | • For both MIC and LIC groups: depression scores decreased over time, adherence levels on HBP treatment recommendations improved, and small increases in high blood pressure knowledge.<br>• Perceived self-efficacy was slightly increased over time in the MIC group and was slightly decreased in the LIC group. | 6/10 |
| Moreno 2012 | USA | • Mental health (depression)<br>• Community health center(s) | • To compare the effectiveness for Hispanic patients of depression treatment provided by a psychiatrist through Internet videoconferencing and treatment as usual by a primary care provider. | • Overall n=182<br>• All Hispanic<br>• Usual care n=87<br>• Intervention=80 | • Telepsychiatry consultations with bilingual providers<br>• Six monthly sessions over 3 years | • The Webcam intervention led to significant improvements in depression severity, functional ability, and quality of life.<br>• The remission rate for patients in the Webcam intervention was also higher (p=.06) | 7/10 |
| Vahia 2015 | USA | • Mental health<br>• Community health center(s) | • To assessed whether neuro-cognitive assessment via telepsychiatry (TP) would be comparable to in-person (IP) testing in a sample of Spanish- speaking older adults in a rural setting. | • Total n=27<br>• All Latino<br>• In-person testing n=11<br>• Telephone testing n=11 | • Telepsychiatry for neuro-cognitive assessment with bilingual providers | • No significant differences between control and intervention test performance, though both groups scored non-significantly higher at the second visit.<br>• This study demonstrates feasibility and utility of neurocognitive testing in Spanish using telepsychiatry among older rural Latinos. | 6/10 |
| Warren 2015 | United Kingdom | Primary care<br>Primary care clinics | To investigate associations between trial patients' demographic, health, and lifestyle characteristics, and their reported experiences of care. | Total n=12,132<br>White 11276, other ethnic =476<br>Usual care =4093<br>GP triage n=4034<br>Nurse triage n=3704 | Telephone triage using computer-supported decision-making software by doctor or nurse. | Compared with white patients, patients from ethnic minorities reported lower satisfaction in all three groups.<br>Patients from ethnic minorities reported higher satisfaction in the GP triage than in usual care<br>Patients from ethnic minorities reported relatively greater ease of getting medical help/advice in the GP triage arm (versus usual care) | 3/10 |
| Yeung 2016 | USA | Mental health<br>Primary care clinics | To evaluate the effectiveness of a telepsychiatry-based culturally sensitive collaborative treatment intervention to improve outcomes for depressed Chinese American immigrants. | Total n=190<br>All Chinese Americans<br>Usual care n=97<br>Telepsychiatry-based culturally sensitive collaborative treatment (T-CSCT) n=93 | Telepsychiatry consultations with bilingual providers | Patients in the T-CSCT group displayed significantly greater improvement in depressive symptoms compared to patients who received usual care | 7/10 |
| **Cohort** |  |  |  |  |  |  |  |
| Berg 2009 | USA | Diabetes<br>Medicaid health plan |  | Total n=980<br>All Hispanic<br>Usual care n=490<br>Intervention n=490 | Disease management diabetes program<br>Scheduled nurse education sessions<br>24-hour access to a nurse counselor | Significant reductions in inpatient bed days for intervention group<br>Significant higher rate of ACE inhibitor and diuretic use for intervention group<br>24.2% reduction in the costs of care. | 10/11 |
| **Quasi-experimental** |  |  |  |  |  |  |  |
| Jang 2014 | USA | • Mental health (depression)<br>• Housing facility | • To explore the feasibility and preliminary efficacy of a telecounseling program in the client's native language. | • n=14<br>• All Korean | • Video-telecounseling consultations with bilingual providers<br>• 4 weekly sessions (30min in each) | • A high level of completion (86%)<br>• High overall satisfaction with the program.<br>• Participants exhibited a significant reduction in depressive symptom severity shortly after completion and at 3-month follow-up. | 5/9 |
| Mayes 2010 | USA | • Diabetes<br>• Community health centers | • To assess the value of using telecommunications with Promotoras (paraprofessional outreach workers) and a medical team in a tele-medicine feasibility project. | • n=16<br>• All Hispanic | • Video-telehealth consultations assisted by Promotoras (bilingual) and primary care physician connecting to a medical specialist | • There was no deterioration in blood pressure control, a modest increase in body weight and a significant decrease in HbA1c. | 4/9 |
| **Cross-sectional** |  |  |  |  |  |  |  |
| Mucic 2010 | Denmark, Sweden | • Mental health<br>• Psychiatric centers and hospitals | • The aim of the present study was to investigate patient satisfaction of a telepsychiatry pilot project. | • n=61<br>• Refugees n=45, asylum seekers (n=12), migrants (n=3), domestic (n=1) | • Video-telepsychiatry consultations with bilingual providers | • Patients reported a high level of acceptance and satisfaction<br>• Patients were willing to use it again or recommend it to others.<br>• Disadvantages of were related to the physical environment | 4/8 |
| Park 2019 | Korea, Vietnam, Cambodia, Uzbekistan | • Health counselling<br>• Digital healthcare centers | • To describe our implementation of the telehealth counseling system and demonstrate the utilization pattern, feasibility, and acceptability of telehealth counseling programs for Korean expatriates. | • n=442<br>• All Korean | • Video-telehealth counseling with bilingual providers | • Overall user satisfaction rates were 96.1%. | 5/8 |
| Schulz 2014 | Australia | • Infectious diseases (e.g. hepatitis C, latent tuberculosis)<br>• Hospital outpatient clinic | • To assess the demographic and disease profile of refugee patients attending a new telehealth clinic, and calculate the patient travel avoided. | • n=119 (patients)<br>• n=29 GPs<br>• Patients: all refugees | • Video-telehealth consultations | • Median distance patient travel distance saved for was 54,000km in total<br>• Technical difficulties were experienced in 25% of consultations | 2/8 |
| Yeung 2009 | USA | • Mental health<br>• Nursing home | • To investigate the feasibility and effectiveness of providing telepsychiatry services to Chinese immigrants in a nursing home | • n=9<br>• All Chinese | • Video-telehealth consultations with bilingual providers | • It was feasible to use videoconferencing to provide follow-up interviews for 8 of the 9 enrolled subjects. | 5/8 |
| **Case series** |  |  |  |  |  |  |  |
| Stewart 2017 | USA | • Mental health<br>• Community trauma treatment centre | • To assess the feasibility of delivering TF-CBT via telehealth using a multiple-case study design | • n=4<br>• All Hispanic | • Weekly Trauma-Focussed CBT (TF-CBT) sessions with bilingual providers | • Telehealth can be a feasible mode to deliver TF-CBT among Hispanic youth. | 7/10 |
| **Mixed methods** |  |  |  |  |  |  |  |
| Bagchi 2018 | USA | • Diabetes, hypertension<br>• Community health center(s) | • To assess the perspectives of consumers and nurses toward a specific telemedicine application. | • n=10<br>• All African American | • Video - telehealth consultations | • Participants viewed the services favourably related to efficiency and convenience.<br>• Few participants said they preferred face-to-face interactions. | 3/7 |
| Glueckauf 2012 | USA | • Mental health (depression)<br>• Health-Care Memory Disorder Clinics | • To assess caregivers' appraisals of the intervention process and to conduct a preliminary analysis of the effects of face-to-face and telephone-based CBT on changes in subjective burden, assistance support, depression and health status | • Total n=12<br>• All African American<br>• Face-to-face CBT n=6<br>• Telephone CBT n=6 | • Telephone CBT<br>• Twelve, 1-hr, weekly sessions | • Both telephone-based and face-to-face CBT showed improvements in depression, subjective burden, and assistance support in caregivers of African Americans with dementia. | 5/7 |
| Mucic 2007 | Denmark | • Mental health<br>• Psychiatric clinic | • To provide psychiatric service on patients' own language where the access to ethnic clinicians is still limited | • Patients n=23<br>• Patients' countries of origin: ex-Yugoslavia, Somalia, Lebanon, Syria<br>• Psychiatrists n= not reported | • Video-telepsychiatry consultations | • Patients reported a high level of acceptance and satisfaction<br>• Patients were willing to use it again or recommend it to others.<br>• Specialists showed general acceptance of telepsychiatry | 0/7 |
| Stewart 2017 | USA | • Mental health<br>• Trauma treatment centre | • A proof-of-concept pilot study of Trauma-Focused CBT (TF-CBT) delivered to underserved trauma-exposed youth and their nonoffending caregivers via telehealth technology | • n=15<br>• Hispanic n=7 (46.7%), African American n=6 (40.0%), Caucasian n=2 (13.3%) | • TF- CBT via video-telehealth with bilingual providers<br>• Weekly sessions (range 12-19 in total) | • Clinically meaningful symptom change post-treatment for youth-reported and caregiver-reported reduction in posttraumatic stress disorder symptoms.<br>• Overall, caregivers were satisfied with telehealth 100% of the time, and 86% | 3/7 |
|  |  |  |  |  |  | indicated that the telehealth equipment was easy to use. |  |
| Ye 2012 | USA | • Mental health<br>• Community health center(s) | • To examine the extent to which the Korean patients accept the telepsychiatry services | • n=16<br>• All Korean | • Video-telehealth consultations with bilingual providers | • Overall, patients felt comfortable during the video interview and were satisfied with the ease of using the teleconferencing equipment.<br>• Having adequate infrastructure to facilitate teleconferencing was important | 4/7 |
| Uebelacker 2011 | USA | • Mental health (depression)<br>• Primary care clinics | • To provide a preliminary test of feasibility, acceptability, and efficacy of telephone depression care management among Latino Medicaid health plan members. | • Total n=38<br>• All Latino<br>• Usual care n=19<br>• Intervention n=19 | • Telehealth depression care management with bilingual providers | • Most participants expressed satisfaction with the program.<br>• There was a trend for the intervention group to experience less depression in time, compared with usual care<br>• Engaging participants via telephone was challenging including numbers out of service. | 6/7 |
| **Qualitative** |  |  |  |  |  |  |  |
| Aisenberg 2012 | USA | • Mental health (depression)<br>• Community health center(s) | • To describe the processes of developing a community-academic partnership that implemented and pilot tested an evidence-based telephone CBT program | • Overall n=101<br>• All Latino<br>• Usual care n=511<br>• Intervention n=50<br>• Physicians n=5 | • Telephone CBT with bilingual providers<br>• Eight-sessions | • Overall, most participants reported very good experiences with the telephone-based delivery of CBT.<br>• Providers expressed strong satisfaction with the participation and retention of clients and the marked<br>improvement of their patients in a relatively short period of time. | 5/10 |
| Pekmezaris 2016 | USA | Heart failure<br>Community | To describes the results of a community based participatory research approach to adapting a telemonitoring HF intervention so that it is acceptable and feasible for use. | n=4<br>African American and Hispanic | Video-telemonitoring consultations<br>Weekly for 3 months | Two major themes related to: 1) suggested changes to the equipment that would maximize usability, 2) suggested changes to the RCT study structure in order to maximize participant engagement. | 5/10 |
| Saberi 2013 | USA | Human immunodeficiency virus (HIV)<br>HIV clinic(s) | To assess the feasibility and acceptability of a telehealth (remote videoconferencing) medication counseling intervention | n=14<br>All African American | Video-telehealth consultations | Participants considered the use of telehealth as a convenient method that has the potential to reduce missed clinic visits due to travel, cost, or privacy concerns. | 5/10 |

**Table 2.** Bias appraisal randomised controlled trial design studies (n=11) (Critical Appraisal Checklist for RCTs (JBI))

| First author<br>(year of<br>publication<br>) | Random<br>-<br>isation<br>reported | Allocati<br>on to<br>groups<br>was<br>conceale<br>d | Groups<br>similar<br>at<br>baseline | Participants<br>blind to<br>group<br>assignment | Those<br>delivering<br>treatment<br>blind to<br>treatment<br>assignment | Outcomes'<br>assessors<br>blind to<br>treatment<br>assignment | Groups treated<br>identically<br>(other than<br>intervention) | Follow-up<br>was<br>complete (for<br>at least 70%<br>of random<br>sample) | Outcome<br>measures<br>reliable,<br>measured<br>similarly<br>for all<br>groups | Appropri<br>ate statistica<br>l<br>analysis<br>used? | Total<br>number of<br>quality<br>indicators<br>reported |
| --- | --- | --- | --- | --- | --- | --- | --- | --- | --- | --- | --- |
| Alcantara<br>(2016)<sup>#</sup> | √ | Not<br>reported | √ | Not reported | Not<br>reported | Not reported | √ | Not reported<sup>*</sup> | √ | √ | 5/10 |
| Chong<br>(2012)<sup>@</sup> | √ | √ | √ | Not reported | Not<br>reported | Not reported | √ | √<sup>*</sup> | √ | √ | 7/10 |
| Dwight-Johnson<br>(2011)<sup>#</sup> | √ | Not<br>reported | √ | Not reported | Not<br>reported | √ | √ | √<sup>*</sup> | √ | √ | 7/10 |
| Han (2010) | √ | Not<br>reported | √ | Not reported | Not<br>reported | Not reported | √ | √<sup>*</sup> | Not<br>reported | √ | 5/10 |
| Himelhoch<br>(2013) | √ | Not<br>reported | √ | Not reported | X | √ | √ | √<sup>*</sup> | √ | √ | 7/10 |
| Jackson<br>(2012)<sup>#</sup> | √ | √ | X | Not reported | Not<br>reported | √ | √ | √<sup>*</sup> | √ | √ | 7/10 |
| Kim<br>(2011)<sup>#</sup> | √ | Not<br>reported | √ | Not reported | Not<br>reported | Not reported | √ | √ | √ | √ | 6/10 |
| Moreno<br>(2012)<sup>@</sup> | √ | √ | √ | Not reported | Not<br>reported | Not reported | √ | √<sup>*</sup> | √ | √ | 7/10 |
| Vahia<br>(2015) | √ | Not<br>reported | √ | Not reported | Not<br>reported | Not reported | √ | √ | √ | √ | 6/10 |
| Warren<br>(2015)<sup>#</sup> | √ | X | X | X | X | X | Not reported | X<sup>*</sup> | √ | √ | 3/10 |
| Yeung<br>(2016) | √ | Not<br>reported | √ | Not reported | Not<br>reported | √ | √ | √<sup>*</sup> | √ | √ | 7/10 |

√ = Yes; X=No; Not reported = absent or insufficient information reported in publication; \*Reported intent to treat approach; # Information re methods supplemented by material re same data set reported in other publications; @ These papers report on the same data set.

**Table 3.** Quality appraisal quasi-experimental design studies (n=2) (Critical Appraisal Checklist for Quasi-Experimental Studies, JBI)

| First author (year of publication) | There is no confusion about which variable comes first? | Participants included in any comparisons were similar? | Participants included in any comparisons received similar treatment (Except for intervention) | There was a control group? | There were multiple measurements of the outcome pre and post the intervention? | Follow up was complete or differences between groups re follow up were adequately reported? | Were the outcomes of participants included in any comparisons measured in the same way? | Were outcomes measured in a reliable way? | Was appropriate statistical analysis used? | Total number of quality indicators reported |
| --- | --- | --- | --- | --- | --- | --- | --- | --- | --- | --- |
| Jang (2014) | √ | N/A* | N/A* | X | √ | √ | N/A* | √ | √ | 5/9 |
| Mayes (2010) | Not reported | N/A* | N/A* | X | √ | √ | N/A* | √ | √ | 4/9 |

√ = Yes; X=No; Not reported = absent or insufficient information reported in publication; N/A=not applicable; \*Single group pre/post design; # Control group noted but nil description re participant selection, recruitment, data collection, intervention, some demographic data comparisons presented at baseline

**Table 4.**
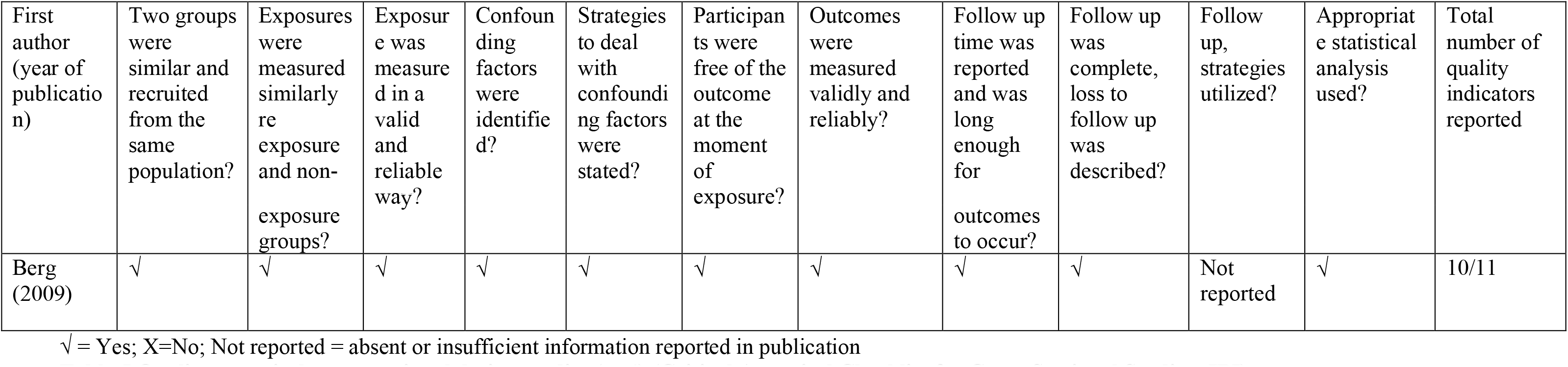
Quality appraisal cohort design studies (n=1) (Critical Appraisal Checklist for Cohort Studies, JBI)

| First author (year of publication) | Two groups were similar and recruited from the same population? | Exposures were measured similarly re exposure and non- | Exposure was measured in a valid and reliable way? | Confounding factors were identified? | Strategies to deal with confounding factors were stated? | Participants were free of the outcome at the moment of exposure? | Outcomes were measured validly and reliably? | Follow up time was reported and was long enough for | Follow up was complete, loss to follow up was described? | Follow up, strategies utilized? | Appropriate statistical analysis used? | Total number of quality indicators reported |
| --- | --- | --- | --- | --- | --- | --- | --- | --- | --- | --- | --- | --- |
|  |  | exposure groups? |  |  |  |  |  | outcomes to occur? |  |  |  |  |
| Berg (2009) | √ | √ | √ | √ | √ | √ | √ | √ | √ | Not reported | √ | 10/11 |

√ = Yes; X=No; Not reported = absent or insufficient information reported in publication

**Table 5.** Quality appraisal cross-sectional design studies (n=4) (Critical Appraisal Checklist for Cross-Sectional Studies, JBI)

| First author (year of publication) | Inclusion criteria were clearly defined? | Participants and setting were described in detail? | Exposure was measured validly and reliably? | Objective, standard criteria used to measure condition? | Confounding factors were identified? | Strategies re confounding factors were stated? | Outcomes were measured validly and reliably? | Appropriate statistical analysis used? | Total number of quality indicators reported |
| --- | --- | --- | --- | --- | --- | --- | --- | --- | --- |
| Mucic (2010) | √ | Not reported | √ | Not reported | Not reported | Not reported | √ | √ | 4/8 |
| Park (2019) | √ | √ | √ | Not reported | Not reported | Not reported | √ | √ | 5/8 |
| Schulz (2014) | Not reported | Not reported | Not reported | Not reported | Not reported | Not reported | √ | √ | 2/8 |
| Yeung (2009) | √ | √ | √ | √ | Not reported | Not reported | Not reported | √ | 5/8 |

√ = Yes; X=No; Not reported = absent or insufficient information reported in publication

**Table 6.** Quality appraisal case series (n=1) (Critical Appraisal Checklist for case series (JBI))

| First author (year of publication) | Clear inclusion criteria? | Condition measured in standardised and | Valid method used to identify condition for all | Case series had consecutive inclusion of participants? | Case series had complete inclusion of participant? | Demographics are clearly reported? | Clinical information is clearly reported? | Follow up results are clearly reported? | Presenting site(s) demographic information | Appropriate statistical analysis used? | Total number of quality indicators reported |
| --- | --- | --- | --- | --- | --- | --- | --- | --- | --- | --- | --- |
|  |  | reliable way? | participants? |  |  |  |  |  | is clearly reported? |  |  |
| Stewart (2017) | √ | √ | √ | Not reported | Not reported | √ | √ | √ | X | √ | 7/10 |

√ = Yes; X=No; Not reported = absent or insufficient information reported in publication

**Table 7.** Quality appraisal mixed methods studies (n=6) (Mixed Methods Appraisal Tool, McGill University)

| First author (year of publication) | Research questions are clear? | Collected data address research questions? | Adequate rationale for mixed methods design? | Different study components are integrated? | Qualitative and quantitative findings are adequately interpreted? | Divergences between quantitative and qualitative results are adequately addressed? | Different components adhere to the quality criteria of each tradition of the methods? | Total number of quality indicators reported |
| --- | --- | --- | --- | --- | --- | --- | --- | --- |
| Bagchi (2018) | √ | √ | Not reported | Not reported | √ | Not reported | Not reported | 3/7 |
| Glueckauf (2012) | √ | √ | √ | √ | √ | Not reported | Not reported | 5/7 |
| Mucic (2007) | Not reported | Not reported | X | X | X | Not reported | Not reported | 0/7 |
| Stewart (2017) | √ | √ | X | X | √ | Not reported | Not reported | 3/7 |
| Uebelacker (2011) | √ | √ | √ | √ | Not reported | √ | √ | 6/7 |
| Ye (2012) | √ | √ | √ | √ | Not reported | X | Not reported | 4/7 |

√ = Yes; X=No; Not reported = absent or insufficient information reported in publication

**Table 8.** Quality appraisal qualitative design studies (n=3) (Critical Appraisal Checklist for Qualitative Studies, JBI)

|  | Congruity re |  |  |  |  |  |  |  |  |  |  |
| --- | --- | --- | --- | --- | --- | --- | --- | --- | --- | --- | --- |
| First author<br>(year of publication) | Philosophical perspective and methodology? | Methodology and the research question/objectives? | Methodology and data collection methods? | Methodology, representation and analysis of data? | Methodology and interpretation of results? | Researcher locates themselves culturally or theoretically? | Influence of researcher on the research is addressed? | Participants' voices are adequately represented? | Ethics approval? | Conclusions are linked to analysis of data? | Total number of quality indicators reported |
| Aisenberg (2012) | Not reported | √ | √ | Not reported | √ | Not reported | Not reported | Not reported | √ | √ | 5/10 |
| Pekmezaris (2016) | √ | Not reported | √ | Not reported | Not reported | Not reported | Not reported | √ | √ | √ | 5/10 |
| Saberi (2013) | Not reported | √ | √ | √ | √ | Not reported | Not reported | X | Not reported | √ | 5/10 |

√ = Yes; X=No; Not reported = absent or insufficient information reported in publication

